# Diagnostic Value Of Turbid Urine In Urogenital Schistosomiasis In Rural Chad Affected By Climate Change Effects: Case Of The Lake Chad Region

**DOI:** 10.1101/2025.05.27.25328414

**Authors:** Didier Lalaye, Tom de Jong, Mirjam de Bruijn, Marius Madjissem, Nan-Arabé Lodoum

## Abstract

**Introduction:** Urogenital schistosomiasis remains a public health issue in sub-Saharan Africa. Diagnosis traditionally relies on detecting eggs in urine or identifying haematuria. The clinical relevance of urine turbidity remains poorly studied.

**Objective:** To assess the relevance of the macroscopic appearance of urine, particularly turbidity, as a presumptive indicator of urogenital schistosomiasis.

**Methods:** A cross-sectional study was conducted with 299 participants in Ngouri, Chad. Urine appearance was visually assessed and compared with urine dipstick results for haematuria detection.

**Results:** Cloudy urine was observed in 20.4% of samples. Among these, 95.1% tested positive on the dipstick, compared to only 16.9% of samples with normal appearance (p < 0.001). Cloudy urine thus appears to be a strong predictive marker.

**Conclusion:** In low-resource settings, visual urine assessment combined with symptom analysis can help guide screening and support presumptive treatment. However, the lack of parasitological confirmation remains a significant limitation.

## INTRODUCTION

Urogenital schistosomiasis is a chronic parasitic disease caused by *Schistosoma haematobium*, a trematode that lodges in the venous plexuses of the urogenital system. Freshwater snails of the genus *Bulinus* host the cercariae, the larval stage that infects humans through skin contact with contaminated water. After migrating to target organs, the adult worms settle in the peri vesical veins, where females lay eggs that pass through the bladder wall, causing local inflammation, urothelial lesions, and potentially severe complications such as fibrosis, hydronephrosis, or the development of bladder carcinomas(1).

The most common clinical manifestations include haematuria, suprapubic pain, urinary disturbances, and cloudy or discoloured urine, often reported by patients. Diagnosis is typically confirmed by identifying eggs in the urine or using dipsticks to detect microscopic hematuria(2).

In tropical and subtropical regions, such as the Lake Chad Basin, schistosomiasis remains a major public health concern, primarily affecting vulnerable rural populations. This vulnerability is exacerbated by climate change, whose effects—rising temperatures, altered water patterns, declining groundwater levels, and the spread of stagnant water bodies—create environmental conditions favourable to the survival and proliferation of snail vectors. These ecological changes contribute to the expansion of transmission zones, increasing the risk of exposure, particularly among children(3).

In our previous studies, we observed that some individuals with cloudy urine, but without visible haematuria, still tested positive on urine dipsticks. This raises the hypothesis that the macroscopic appearance of urine could serve as a useful clinical indicator, complementing standard screening methods.

The present study therefore aims to explore the relationship between urine appearance, dipstick results, and the potential presence of schistosomiasis, with the goal of improving field screening strategies in a context increasingly challenged by logistical, health, and climate-related constraints.

### RATIONALE FOR THE STUDY

In resource-limited settings, schistosomiasis diagnosis often relies on accessible methods, such as symptom assessment and/or urine dipstick tests. Our field observations showed that some samples of cloudy urine, even without visible haematuria, were associated with positive dipstick results. This suggests that visual assessment could serve as a useful preliminary indicator, particularly for community health workers.

At the same time, climate change—through rising temperatures, changing rainfall patterns, and the formation of stagnant water bodies—is altering snail habitats and expanding schistosomiasis transmission zones(4). This evolving situation calls for increased vigilance and the optimization of simple, rapid detection tools.

In this context, it becomes relevant to systematically assess the diagnostic value of urine appearance in relation to dipstick test results, in order to improve early case identification in remote areas

### HYPOTHESES

Null hypothesis (H□): There is no significant association between the macroscopic appearance of urine and positivity on urine screening tests for urogenital schistosomiasis.

Alternative hypothesis (H□): There is a significant association between the macroscopic appearance of urine and positivity on urine screening tests for urogenital schistosomiasis.

### OBJECTIVES

#### General objective

To evaluate the diagnostic value of the macroscopic appearance of urine as a presumptive indicator of urogenital schistosomiasis in a rural endemic area of Lake Chad, by examining its association with haematuria detected by urine dipstick tests.

#### Specific objectives

- To determine the prevalence of urogenital schistosomiasis according to the macroscopic appearance of urine and assess the strength of this association.
- To compare the frequency of positive tests between urine samples with normal appearance and those showing visible abnormalities (cloudiness or haematuria).
- To examine the relationship between reported symptoms (urinary disturbances, abdominal pain) and urine appearance.

## MATERIALS AND METHODS

### Sampling and Study Design

This is a cross-sectional, descriptive, and prospective study conducted from July 1 to July 15, 2024, in the Ngouri district, Chad. The data collection used the Dawa Mobile Health system, which was previously effective in Torrock for detecting and managing urogenital schistosomiasis(5).

The survey was conducted across several villages under the jurisdiction of the Ngouri district hospital. Participants included adult volunteers as well as children whose parents requested screening. A total of 300 individuals were enrolled through simple random sampling within each village. One case with incomplete data was excluded, resulting in a final sample size of 299 participants.

Each participant was assigned an identification code by a local healthcare worker, which was recorded in a registry. Sociodemographic and clinical data were collected using a standardized questionnaire, and a urine sample was obtained from each subject.

### Study Area (Fig. 1)

This study was conducted in the Ngouri sub-prefecture, located in the Lac province in western Chad, approximately 230 km north of N’Djamena. This agro-pastoral region covers an area of 3,375 km^2^ and has an estimated population of 262,250 inhabitants, with a sex ratio of 0.98 (https://fr.city-facts.com/ngouri-td/population).

**Fig 1:**
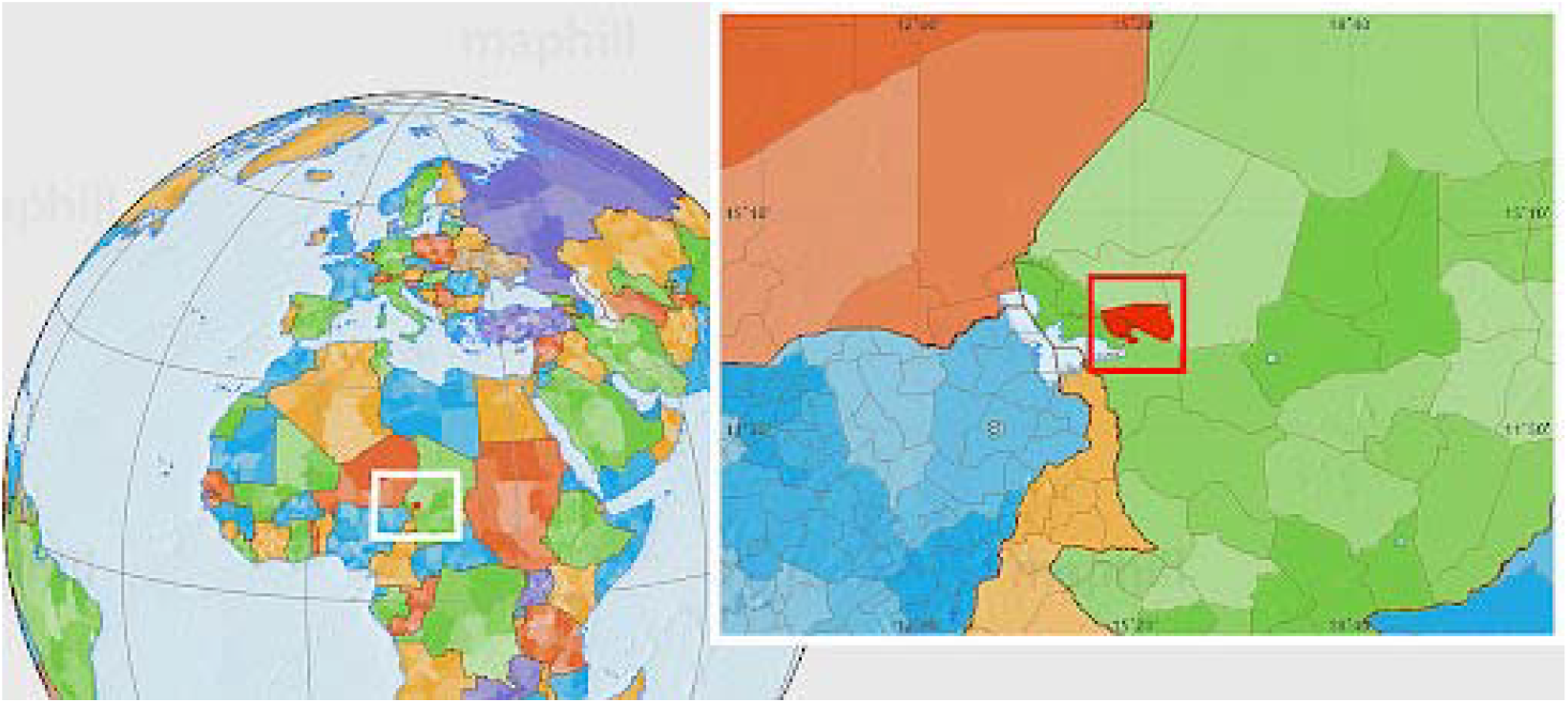
Map of the Ngouri District (source: Maphill)

**Fig. 2:**
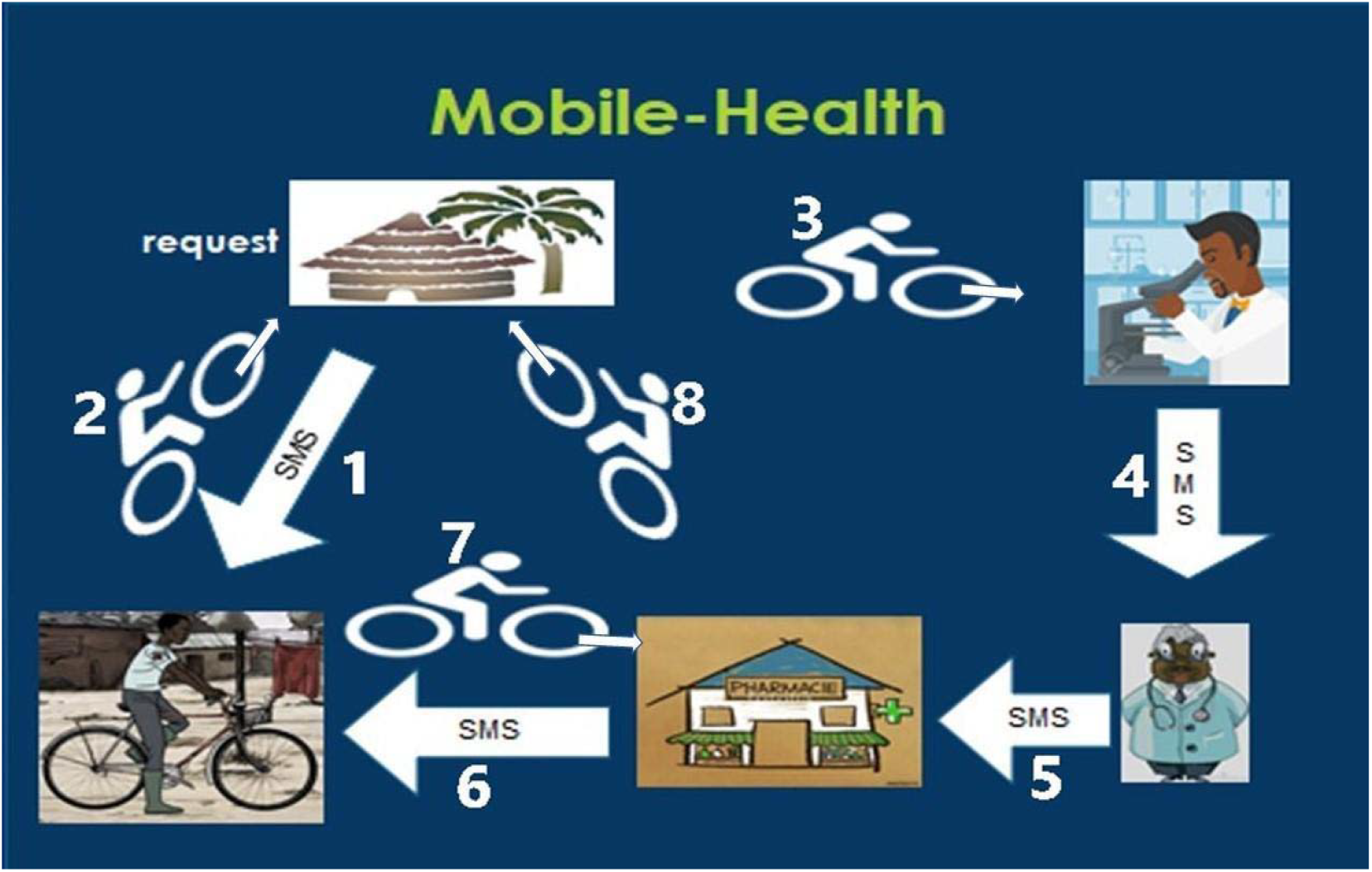
Operation of the Mobile Health System.

**Fig 3:**
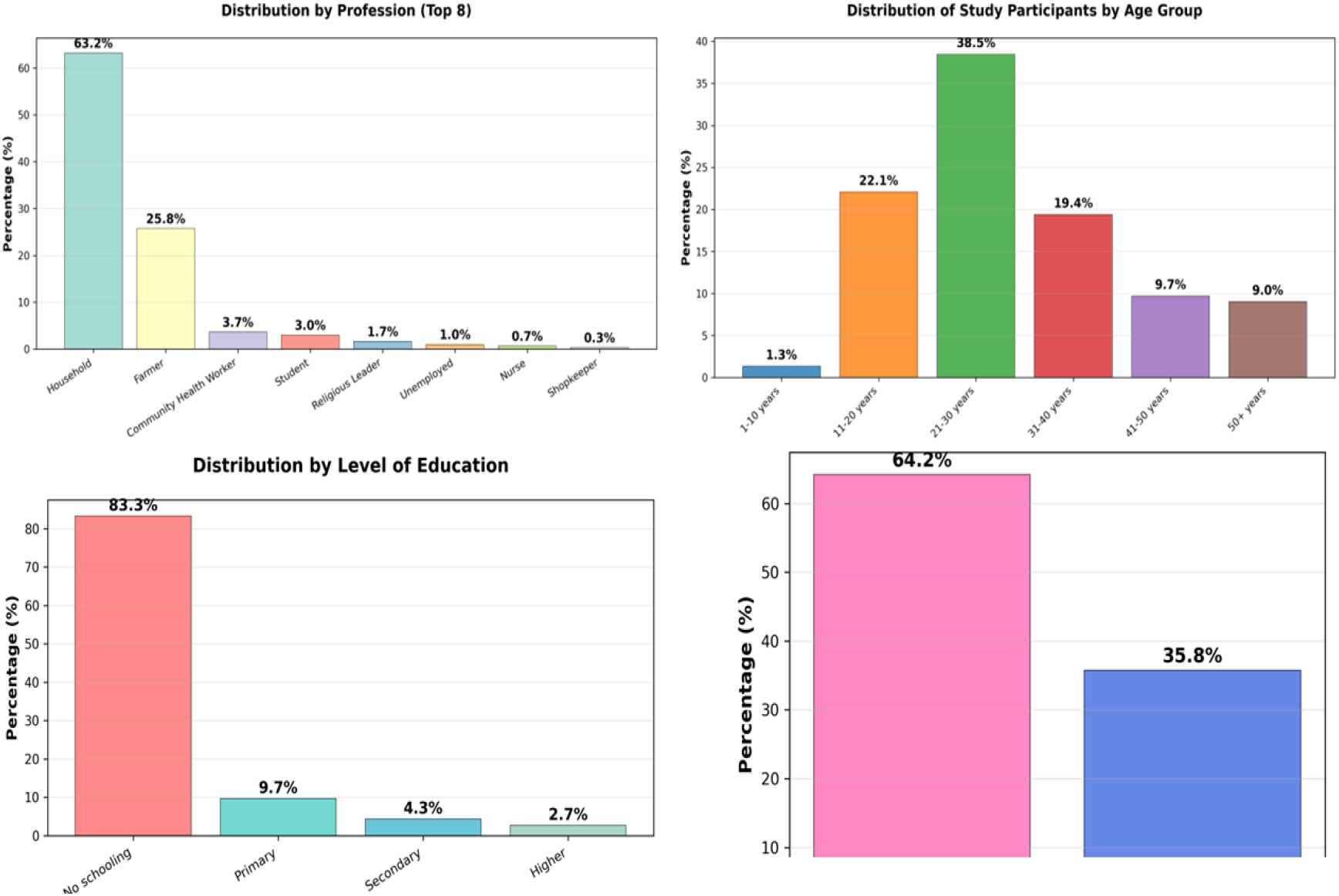
Proportion of study participants by sociodemographic characteristics. Most participants (38.5%) were aged between 21 and 30 years. The age groups 11–20 years (22.1%) and 31–40 years (19.4%) were well represented. The mean age was 29.6 years (± standard deviation: 12.2 years).

**Figure 4:**
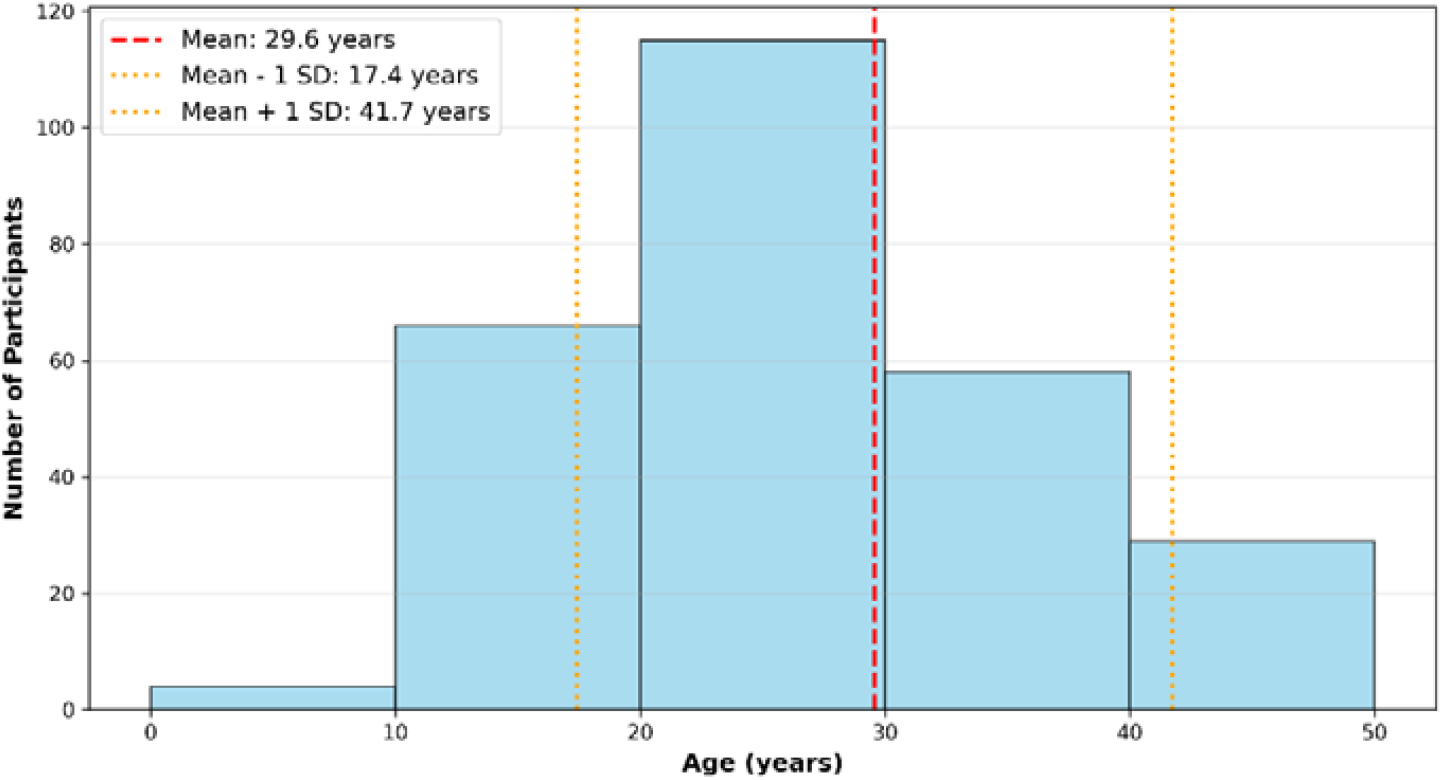
Age distribution of study participants, including mean and standard deviation. The study population consisted of 64.2% women and 35.8% men, yielding a sex ratio of 0.56 male to female.

**Fig 5:**
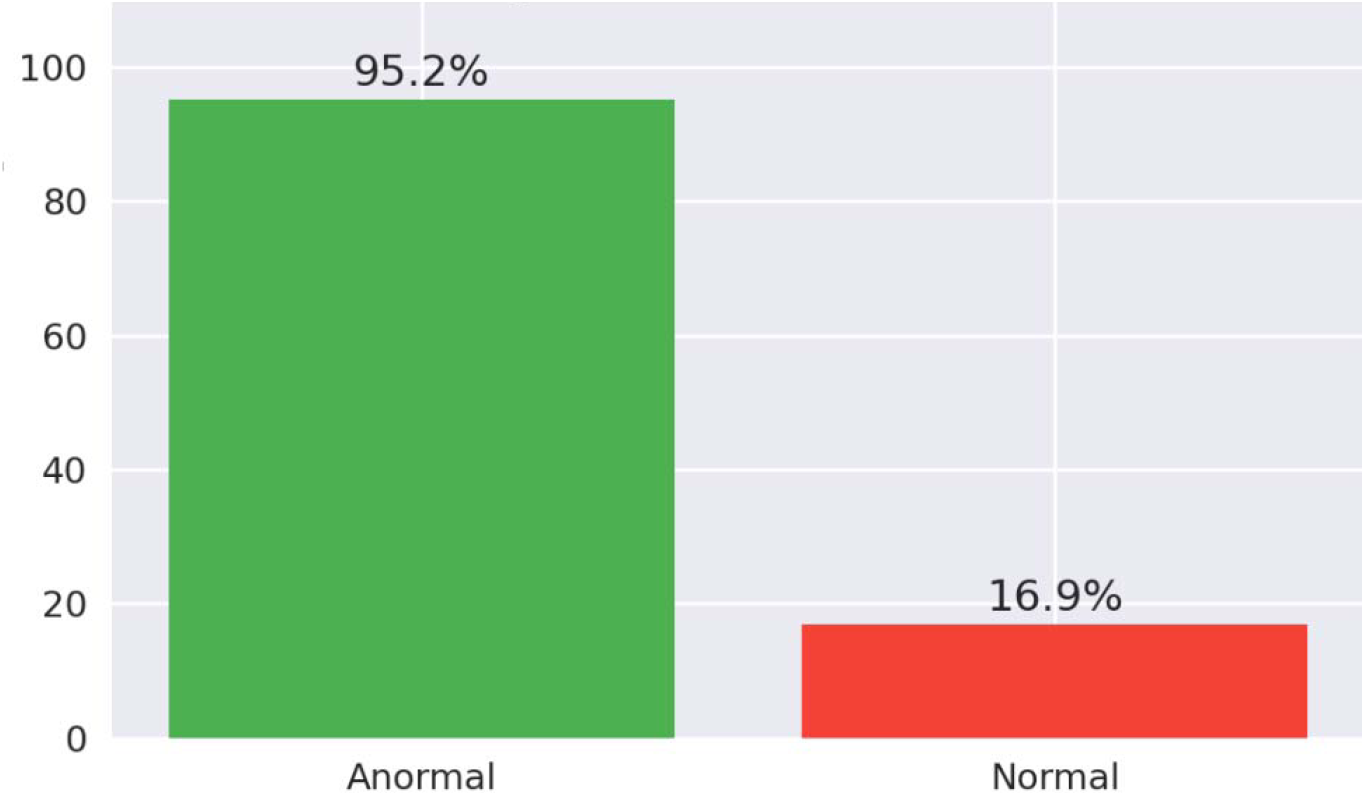
Frequency of positive dipstick results according to urine appearance (normal vs abnormal)

**Fig 6:**
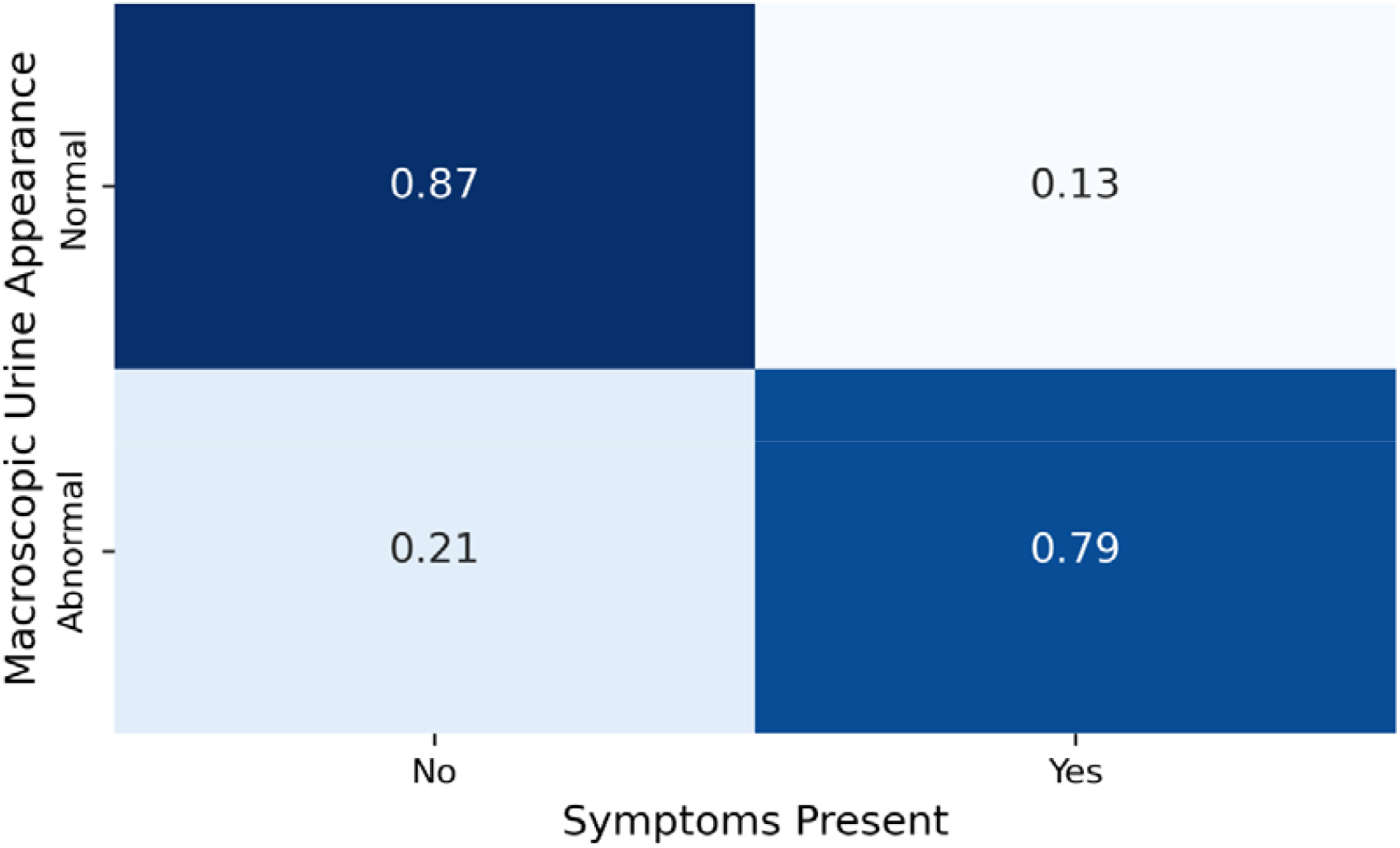
Proportion of symptoms by urine appearance. Participants with normal urine appearance were mostly asymptomatic (87.3%), with only 12.7% reporting symptoms. In contrast, 79.0% of those with abnormal urine appearance had symptoms. This association is highly significant (p<0.001*p*<0.001), and the odds of having symptoms are 26 times higher for those with abnormal urine.

The Ngouri Health District, one of the seven operational districts within the Provincial Health Delegation of Lac, was selected due to its high exposure to urogenital schistosomiasis. The progressive drying of Lake Chad, driven by climate change, has led to the emergence of stagnant water bodies in surrounding villages, creating favourable conditions for the proliferation of *Schistosoma haematobium*.

The district is bordered by:

- Mao Health District (Kanem province) to the northwest;
- Kouloudia Health District to the south;
- Mondo (Kanem) and Massakory (Hadjer Lamis) Health Districts to the east;
- Dibinintchi and Isseirom Health Districts to the west.

From a healthcare perspective, the area is served by 20 health centres, most of which lack laboratories and qualified personnel, as well as a district hospital located in Ngouri. To date, no specific study on urogenital schistosomiasis has been conducted in this locality, and no mass treatment strategy has yet been implemented.

### Communication

The community was informed about the availability of urogenital schistosomiasis testing through urine sampling at home, regardless of symptom presence. This information was disseminated via multiple channels, including churches, mosques, markets, schools, and local FM radio stations, in Kanembou (the local language), Arabic, and French.

### Diagnostic and Treatment Process

Urine samples were collected between 9 a.m. and 1 p.m., and children participating in the study were weighed. Samples were transported to the laboratory within three hours of collection. Only samples testing positive for haematuria by urine dipstick were considered positive cases and were treated free of charge with praziquantel provided by the regional hospital.

The mobile health system workflow operates as follows:

- A hospital agent travels to villages to collect urine samples and transport them to the hospital laboratory for analysis. In case of a positive diagnosis, the agent ensures the delivery of praziquantel.
- The laboratory technician analyses the urine samples and notifies the physician of positive schistosomiasis cases.
- The physician receives the test results via SMS, calculates the praziquantel dosage based on the child’s weight, and sends the prescriptions to the pharmacist via SMS.
- The pharmacist is responsible for dispensing the medication, labelled with the patient’s name and personal code, and informs the field agent by SMS for delivery to the families.

### Data Analysis

Data were collected on printed forms, entered into Excel, and analysed with SPSS version 25 (IBM SPSS Statistics, New York, NY). All association tests were conducted with a significance threshold set at 5% (p < 0.05).

To investigate the existence of a statistical association between the macroscopic appearance of urine and dipstick positivity, the Chi-square test of independence was applied, with significance set at p < 0.05.

### Inclusion Criteria

All residents of the Ngouri sub-prefecture who consented to participate in the study for adults, and children over one year of age whose parents gave consent.

### Ethical Considerations

Written authorization was obtained from the Ngouri district health authority. Oral consent was obtained from adult participants and parents of minor participants prior to sample collection. Each participant or parent signed a dedicated column in the register. Collected data were anonymized before analysis. An exemption from ethical approval was granted by the National Bioethics Committee of Chad.

## RESULTS

### Sociodemographic Characteristics Of The Study Population

A large majority of respondents (83.3%) had never attended school. Only 9.7% reached primary education, 4.3% secondary, and 2.7% higher education.

The most common occupation was “housewife” (i.e., women not engaged in any formal employment), representing 63.2%, followed by “farmer” at 25.8%.

### Results of Urine Tests and Association with Macroscopic Urine Appearance

Out of the 299 urine samples analysed, 87 (29.1%) tested positive for haematuria using dipstick tests, while 212 (70.9%) were negative.

Cross-tabulating these results with the urine’s macroscopic appearance revealed a clear pattern. Among samples with abnormal appearance (cloudy or hematic), 95.2% were dipstick-positive, compared to only 16.9% of samples with normal appearance (Table 1). This association was highly significant (p < 0.001), with an odds ratio of 0.01, indicating an extremely low probability of a positive test in the absence of visible urine abnormalities (Figure 5).

**Table 1.** Distribution of dipstick test results according to macroscopic urine appearance (n=299)

| *Urine Appearance* | *Positive Dipstick n (%)* | *Negative Dipstick n (%)* | *Total n (%)* |
| --- | --- | --- | --- |
| **Normal** | 40 (16.9) | 197 (83.1) | 237 (79.3) |
| **Cloudy (Turbid)** | 58 (95.1) | 3 (4.9) | 61 (20.4) |
| **Hematic** | 1 (100) | 0 (0) | 1 (0.3) |
| **Total** | 87 (29.1) | 200 (70.9) | 299 (100) |

Specifically, cloudy urine samples (n = 61) accounted for 20.4% of the total. Of these, 58 (95.1%) tested positive. The single hematic urine sample also tested positive. Among the 237 samples with normal urine appearance, 40 (16.9%) tested positive on the dipstick test.

These findings confirm that the visual appearance of urine, particularly turbidity, serves as a relevant clinical marker for urogenital schistosomiasis, although a non-negligible proportion of positive cases were also detected among subjects with normal-appearing urine.

### Association Between Macroscopic Urine Appearance And Presence Of Urinary Or Abdominal Symptoms

## DISCUSSION

### Sociodemographic characteristics of the study population

The mean age of participants was 29.6 years (± 12.2), reflecting a predominance of young adults. This finding is consistent with other studies in rural settings: Gessessew Bugssa et al. reported a mean age of 25 years in Alamata (Ethiopia), and Gazzinelli et al. found approximately 30 years in Brazil, both in schistosomiasis-endemic areas(6,7). In contrast, Faust et al. observed a lower average age (9–12 years) during mass drug administration campaigns targeting school-aged children, which explains the difference observed in our more intergenerational, voluntary, community-based approach(8).

Participants aged 21–30 years accounted for 38.5%, and those aged 11–20 years made up 22.1%, which aligns with rural demographic pyramids in developing countries. The high proportion of women (64.2%) may be explained by their greater availability, as noted by Colley et al. in 2014, and by male rural-to-urban migration in search of employment(9,10) The low level of formal education (83.3% with no schooling) represents a well-documented barrier to prevention programs, as emphasized by Ngassa et al., and is worsened by high dropout rates: only 9.7% completed primary education, 4.3% secondary, and 2.7% higher education(11).

Lastly, the occupational distribution, dominated by Households (63.2%) and farmers (25.8%), reflects the local subsistence economy. These groups are particularly exposed to contaminated water sources, the main transmission route for schistosomiasis, as stated by the World Health Organization(12)

#### Diagnostic Value of Urine Appearance, with a Particular Focus on Turbidity

Macroscopic examination of urine represents an accessible and pragmatic tool for guiding the diagnosis of urogenital schistosomiasis, particularly in low-resource community settings. In our study, 79.3% of the urine samples had a normal appearance, 20.4% were turbid, and 0.3% were hematuric. This distribution is comparable to that reported by Senghor et al. (2014) in Senegal(13).

Among turbid urine samples, 95.1% tested positive using urine dipsticks—representing the highest predictive value across all urine appearance categories. This remarkable diagnostic performance is often underestimated in classical approaches focused solely on haematuria. These findings reinforce the observations of Houmsou et al. (2011) in Nigeria, who emphasized the clinical relevance of turbidity as an early warning sign for schistosomiasis(14).

In contrast, although macroscopic haematuria is classically considered pathognomonic, it was extremely rare in our sample (0.3%).

While normal-appearing urine constituted the majority (79.3%), it did not rule out infection: 16.9% of these samples tested positive. This confirms that macroscopic examination alone is insufficient, aligning with the findings of Colley et al., who highlighted the frequency of asymptomatic or mild infections(10).

Statistical analysis confirmed a highly significant association between abnormal appearance (turbid or hematuric) and dipstick positivity (p < 0.001; OR = 0.01). However, our study clearly demonstrates that urine turbidity is an underestimated yet highly reliable indicator that deserves to be emphasized in early detection strategies, particularly in school settings or mobile outreach campaigns.

In total, while haematuria is well-known for guiding diagnosis, turbid urine emerges as an equally relevant—and often more frequent—marker, and thus potentially more useful in field conditions. It should not be considered a secondary sign but rather a major clinical alert, especially where diagnostic resources are limited.

#### Relationship Between Urine Appearance and Symptoms

The strong association between abnormal urine appearance—particularly turbid or hematuric—and the presence of specific clinical symptoms (here grouped under the term *“symptomatic”*, which includes signs such as dysuria, abdominal pain, and pollakiuria) represents a valuable indicator for guiding the diagnosis of urogenital schistosomiasis in field settings.

In practical terms, the simultaneous presence of turbid urine and one or more suggestive symptoms significantly increases the likelihood of infection with *Schistosoma haematobium*. This correlation aligns with findings from several studies on the clinical diagnosis of schistosomiasis, where macroscopic urine inspection combined with symptom assessment effectively identifies individuals at risk(15,16).

In contexts where access to confirmatory laboratory testing is limited—such as in the absence of diagnostic facilities, during mass treatment campaigns, or in resource-constrained settings—this combination of clinical signs may justify the use of presumptive treatment. This approach is endorsed by the World Health Organization in endemic areas, as it helps ensure timely care for likely-infected individuals, thereby reducing the risk of severe complications(12)

Thus, the combination of macroscopic urine examination with basic clinical symptom assessment constitutes a simple, rapid, and relevant tool for medical decision-making in rural or low-resource environments. Where feasible, it can guide referrals for targeted confirmatory testing, optimizing available resources. Otherwise, it provides a solid basis for initiating effective presumptive treatment, contributing to the protection of at-risk populations.

#### Study Limitations

This study is limited by its community-based context and resource constraints. Diagnosis relied exclusively on the detection of haematuria via urine dipsticks, without parasitological confirmation (e.g., microscopy or antigen testing), which prevents definitive identification of *Schistosoma haematobium* and differentiation from other causes of haematuria.

The visual assessment of urine appearance was performed subjectively by field agents, introducing potential variability depending on observation conditions. Additionally, no investigation was conducted for urinary co-infections such as bacterial infections, which may confound the interpretation of observed abnormalities.

Finally, voluntary participation may have led to overrepresentation of individuals already experiencing symptoms, introducing a selection bias.

While these limitations are understandable in an operational field setting, they highlight the need for future studies incorporating more comprehensive diagnostic tools and broader screening for urinary tract conditions.

## CONCLUSIONS

This community-based study in the Lake Chad region highlights the diagnostic value of macroscopic urine examination—particularly urine turbidity—as a practical and reliable indicator for urogenital schistosomiasis. While macroscopic hematuria is widely recognized as a hallmark symptom, the findings of this study emphasize that cloudy urine is a frequent and strongly predictive marker, with a 95.1% positivity rate on dipstick tests. Despite its diagnostic potential, this indicator is often overlooked in conventional screening protocols.

These results underscore the importance of including urine turbidity as a key decision-making criterion in low-resource settings, where access to laboratory diagnostics is limited. At the same time, the presence of positive cases among individuals with normal-appearing urine reinforces the need for complementary tools—such as urine dipstick testing—to improve diagnostic accuracy and avoid missed cases.

In conclusion, the study advocates for a renewed clinical emphasis on urinary turbidity within schistosomiasis control strategies. It supports the integration of simple, syndromic diagnostic approaches that combine visual urine inspection with symptom assessment to expand access to timely diagnosis and treatment in endemic and resource-constrained environments.

Despite the absence of parasitological confirmation, this research offers valuable insights for health system innovation and schistosomiasis control in climate-affected regions. It demonstrates that urinary turbidity can serve as a practical, field-level proxy and should be incorporated into community-based screening and mobile health interventions.

## Data Availability

All data produced in the present study are available upon reasonable request to the authors

## AUTHOR CONTRIBUTIONS

Didier Lalaye conceptualized the study, coordinated field operations, and drafted the manuscript.

Tom de Jong contributed to the study design, performed data analysis, and critically revised the manuscript.

Mirjam de Bruijn, an anthropologist, contributed to the analysis and interpretation of sociodemographic data and reviewed the manuscript for contextual relevance.

Marius Madjissem supported data collection, facilitated field logistics, and participated in community engagement.

Nan-Arabé Lodoum designed the urine sampling protocol and contributed to the correction and validation of the database.

## RÉFÉRENCES

1. Frandsen F. Studies of the relationships between Schistosoma and their intermediate hosts. II. The genus Bulinus and Schistosoma haematobium from Sudan, Zaire and Zambia. J Helminthol. 1979;53(3).

2. Vaillant MT, Philippy F, Neven A, Barré J, Bulaev D, Olliaro PL, et al. Diagnostic tests for human Schistosoma mansoni and Schistosoma haematobium infection: a systematic review and meta-analysis. Lancet Microbe. 2024;5(4).

3. Adekiya TA, Aruleba RT, Oyinloye BE, Okosun KO, Kappo AP. The effect of climate change and the snail-schistosome cycle in transmission and bio-control of schistosomiasis in sub-saharan africa. Vol. 17, International Journal of Environmental Research and Public Health. MDPI AG; 2020.

4. Nwoko OE, Manyangadze T, Chimbari MJ. Predicted changes in habitat suitability for human schistosomiasis intermediate host snails for modelled future climatic conditions in KwaZulu-Natal, South Africa. Front Environ Sci. 2023;11.

5. Lalaye D, de Bruijn ME, de Jong TPVM. Impact of a mobile health system on the suppression of schistosoma haematobium in Chad. American Journal of Tropical Medicine and Hygiene. 2021 Oct 1;105(4):1104–8.

6. Bugssa G, Teklehaymanot T, Medhin G, Berhe N. Prevalence and intensity of Schistosoma mansoni infection, and contributing factors in Alamata district of Tigray Region, Northern Ethiopia. PLoS Negl Trop Dis. 2024 Nov 1;2024-November.

7. Gazzinelli A, Bethony J, Fraga LA, LoVerde PT, Correa-Oliveira R, Kloos H. Exposure to Schistosoma mansoni infection in a rural area of Brazil. I: Water contact. Tropical Medicine and International Health. 2001;6(2).

8. Faust CL, Osakunor DNM, Downs JA, Kayuni S, Stothard JR, Lamberton PHL, et al. Schistosomiasis Control: Leave No Age Group Behind. Vol. 36, Trends in Parasitology. 2020.

9. Tacoli C, Mcgranahan G, Satterthwaite D. WORLD MIGRATION REPORT Urbanization, Rural-urban Migration and Urban Poverty Background paper Background paper Migrants and Cities: New Partnerships to Manage Mobility. 2014.

10. Colley DG, Bustinduy AL, Secor WE, King CH. Human schistosomiasis. The Lancet [Internet]. 2014;383(9936):2253–64. Available from: 10.1016/S0140-6736(13)61949-2

11. Ngassa N, Zacharia A, Lupenza ET, Mushi V, Ngasala B. Urogenital schistosomiasis: prevalence, knowledge and practices among women of reproductive age in Northern Tanzania. IJID Regions. 2023;6.

12. WHO. Weekly epidemiological record: Schistosomiasis and soil-transmitted helminthiases: numbers of people treated in 2017 [Internet]. Vol. 50, Weekly Epidemiological Record. 2018 [cited 2025 May 24]. Available from: https://www.who.int/news-room/fact-sheets/detail/schistosomiasis

13. Senghor B, Diallo A, Sylla SN. Prevalence and intensity ofurinary schistosomiasis among school children in the districtof Niakhar, region of Fatick, Senegal. Parasit Vectors. 2014;7:1–6.

14. Houmsou RS, Amuta EU, Sar TT. Profile of an epidemiological study of urinary schistosomiasis in two local government areas of Benue state, Nigeria. International Journal of Medicine and Biomedical Research [Internet]. 2012;1(1):39–48. Available from: www.ijmbr.com

15. Fernández-Soto P, Avendaño C, Sala-Vizcaíno A, Crego-Vicente B, Febrer-Sendra B, García-Bernalt Diego J, et al. Molecular Markers for Detecting Schistosoma Species by Loop-Mediated Isothermal Amplification. Dis Markers. 2020;2020.

16. Casacuberta-Partal M, Janse JJ, van Schuijlenburg R, de Vries JJC, Erkens MAA, Suijk K, et al. Antigen-based diagnosis of Schistosoma infection in travellers: A prospective study. J Travel Med. 2021;27(4).

